# The association between undetected heteroresistance and antibiotic treatment failure in complicated urinary tract infection

**DOI:** 10.1101/2025.03.11.25323422

**Authors:** Carter N. Abbott, Aditi Dhillon, Sushma Timalsina, Elise Furr, Patrick Velicitat, Adam Belley, Navaneeth Narayanan, Keith S. Kaye, David S. Weiss

**Author notes:** Corresponding authors: David S. Weiss, Emory Antibiotic Resistance Center, 954 Gatewood Rd. Room 2052, Atlanta, GA, 30329., Keith S. Kaye, Department of Medicine, Rutgers Robert Wood Johnson Medical School, New Brunswick, NJ, 08901.

## Abstract

**Background:** Antibiotic resistance is a worsening public health threat. One poorly understood aspect of this problem is unexpected antibiotic treatment failure; when an infecting isolate is deemed susceptible to a given antibiotic, yet treatment with that drug fails. It has been proposed that heteroresistance may be an explanation for at least some unexplained treatment failures. Heteroresistance occurs when a bacterial isolate harbors a minor subpopulation of resistant cells which coexists with a majority susceptible population. The clinical relevance of heteroresistance is not clear.

**Methods:** We obtained 291 index isolates from 288 unique patients in the piperacillin/tazobactam arm of the ALLIUM phase 3 clinical trial for the treatment of Gram-negative pathogens causing complicated urinary tract infections. The MIC for all isolates was below the piperacillin/tazobactam resistance breakpoint according to standard antimicrobial susceptibility testing. We performed population analysis profiles on these isolates to detect piperacillin/tazobactam heteroresistance and conducted a post hoc analysis to examine the impact of heteroresistance on clinical outcomes.

**Findings:** We observed that 33/288 (11.5%) of the patients were infected with isolates that were heteroresistant to piperacillin/tazobactam and that patients infected with heteroresistant isolates had an increased rate of treatment failure when compared to patients infected with a non-heteroresistant isolate (odds ratio [OR] 2.13, 95% CI 1.02, 4.41; adjusted OR 1.74, 95% CI 0.82, 3.71). Further, patients without a removable catheter were at particular risk of treatment failure from infection with heteroresistant isolates.

**Interpretation:** These data demonstrate that patients infected with a piperacillin/tazobactam heteroresistant isolate are at an increased risk for piperacillin/tazobactam treatment failure. The results help contextualize commonly observed unexpected antibiotic treatment failure and highlight heteroresistance as a potential cause.

**Funding:** This work was, in part, funded by the United States Government, Advanced Research Projects Agency for Health (ARPA-H) grant AY1AX000002, National Institutes of Health (NIH) grant AI158080, and Department of Veterans Affairs (VA) grant BX005985 to DSW. DSW was also supported by a Burroughs Wellcome Fund Investigator in the Pathogenesis of Infectious Disease award. NN was supported by the National Institute of Allergy and Infectious Diseases (NIAID) of the National Institutes of Health (NIH) under Award Number K23AI159396. The views and conclusions contained in this document are those of the authors and should not be interpreted as representing the official policies, either expressed or implied, of the United States Government.

**Competing Interests:** PV is an employee of Allecra Therapeutics. AB was a paid consultant for Allecra at the time of the study. NN reports grants/contracts from Merck and Shionogi; consulting/speaker fees from Astellas. KSK has received consulting fees from Merck, Shionogi, GlaxoSmithKline, MicuRx Pharmaceuticals, AbbVie, Venatorx Pharmaceuticals, and Allecra Therapeutics, and owns stock options in Merck. DSW has pending patents related to heteroresistance.

## Introduction

Antimicrobial resistance poses a significant threat to public health. In 2019 alone, approximately 1.27 million deaths were attributed to antimicrobial resistance and this global crisis is expected to worsen (1). Projections indicate that by 2050, antimicrobial resistance could be responsible for 10 million worldwide deaths annually (2). To combat this threat, gaining a better understanding of mechanisms of antibiotic resistance and their contributions to treatment failure could help optimize treatment regimens.

One clinical issue that currently lacks adequate attention is unexpected treatment failure. Unexpected treatment failure occurs when a bacterial isolate infecting a patient is not eradicated despite the isolate appearing susceptible to the given antibiotic based on *in vitro* laboratory testing. One possible cause of treatment failure is heteroresistance, a form of antibiotic resistance where a minor subpopulation of resistant cells co-exists with a majority susceptible population. Notably, these resistant subpopulations are often undetected by standard susceptibility testing methods yet rapidly replicate in the presence of a given antibiotic.

The clinical significance of heteroresistance remains uncertain. Studies employing murine infection models have demonstrated that heteroresistance can mediate treatment failure (3–5). Additionally, there have been documented case studies wherein antibiotic therapy failed to eliminate an infection with an isolate heteroresistant to the antibiotic utilized for treatment (6, 7).

Given the limited clinical data regarding the impact of heteroresistance on treatment outcomes, there remains a critical gap in understanding. Accordingly, we performed a post hoc analysis of the ALLIUM trial to examine the effect of heteroresistance on clinical outcomes amongst complicated urinary tract infection (cUTI) subjects treated with piperacillin/tazobactam. Particular focus was given to cUTI without a removeable source of infection, due the higher treatment failure rate in this type of UTI. We hypothesized that heteroresistance might associated with treatment failure in cUTI without a removeable source of infection.

## Materials and Methods

### Study Design and Population

We obtained clinical isolates from the ALLIUM clinical trial (NCT03687255), a phase 3, randomized, double blinded trial to compare the efficacy of piperacillin/tazobactam versus cefepime/enmetazobactam in treating hospitalized patients with ongoing cUTI, including acute pyelonephritis (AP) (8). In this study, we performed secondary nested cohort analyses, in concert with population analysis profiles, to identify heteroresistant isolates and assess heteroresistance as a potential contributor to clinical outcome. The 291 isolates we obtained from this clinical trial originated from patients assigned to the piperacillin/tazobactam arm, specifically from the microbiologically evaluable subset, and were collected prior to any patient exposure to piperacillin/tazobactam. Each patient received intravenous treatment with piperacillin/tazobactam and was clinically evaluated at day 3 of treatment, end of treatment (EOT), test of cure (TOC; EOT +7 days [+/- 2 days]), and late follow up (LFU; EOT + 14 days [+/- 2 days]). Treatment outcome was determined by a composite of clinical and microbiological outcomes with microbiological cure being defined as a reduction in urine bacterial load to <10^3^ CFU/mL. Additional information pertaining to the trial protocol is available https://classic.clinicaltrials.gov/ct2/show/NCT03687255 (8).

### Clinical Isolates

The study population consisted of 288 patients from whom 291 Gram-negative isolates were obtained. The isolates consisted of 216 *Escherichia coli*, 30 *Klebsiella* spp., 20 *Proteus* spp., 11 *Pseudomonas aeruginosa*, 4 *Enterobacter* spp., 3 *Citrobacter* spp., 3 *Serratia marcescens*, 2 *Acinetobacter ursingii*, 1 *Morganella morganii*, and 1 *Providencia rettgeri* (Supplementary Table 1). The MIC of all isolates included in this study was below the piperacillin/tazobactam resistance breakpoint according to the 2024 CLSI susceptibility interpretive criteria for broth microdilution testing (9). Enterobacterales isolates with MICs at the susceptible-dose dependent breakpoint of 16/4 µg/mL were included due to the extended infusion utilized in the trial (4 g piperacillin, 0.5 g tazobactam, by 2-hour infusion every 8 hours) and because piperacillin/tazobactam is concentrated at high levels in the urine (8). Breakpoints of <64/4 µg/mL and <128/4 ug/mL were used for *P. aeruginosa* and *A. ursingii*, respectively. All isolates were part of the microbiologically evaluable population of the piperacillin/tazobactam arm of the trial (8). The bacterial isolates were acquired from IHMA and stored at -80°C.

### Population Analysis Profile

Population analysis profile (PAP) was performed by inoculating 1.5 mL of Mueller-Hinton Broth (MHB) with a single colony streaked from a frozen stock of a given isolate. The culture was incubated overnight for 16-18 hours in a 37°C shaking incubator at 200 RPM. The culture was then serially diluted in PBS 1:10 over a series of 6 dilutions. The undiluted sample and the dilution series were then plated together on Mueller-Hinton Agar (MHA) plates containing no drug, 4/4, 8/4, 16/4, 32/4, or 64/4 ug/mL of piperacillin/tazobactam (MedChemExpress). Colonies were enumerated after a 15-18 hour incubation at 37°C. Isolates were classified resistant if they exhibited at least 50% survival on the 16/4 ug/mL piperacillin/tazobactam MHA plate as compared to the number of colonies surviving on the MHA plate without drug. Isolates were classified heteroresistant if they had less than 50% survival, but more than 0.0001% survival, on two out of the three piperacillin/tazobactam concentrations (8/4 ug/mL, 16/4 ug/mL, and 32/4 ug/mL) when compared to growth on the MHA plate without drug. Isolates that were not classified resistant or heteroresistant were classified susceptible by PAP, and from this point forward, will be referred to as non-heteroresistant (Supplementary Figure 1). Single biological replicates were used, and each strain was assessed by PAP ≥3 times. In instances where patients had more than one pathogen, if one or more of the pathogens is heteroresistant then the patient’s infection was categorized as heteroresistant; if none were heteroresistant then the infection was categorized as non-heteroresistant.

### Genome analysis

DNA was extracted using the Wizard Genomic DNA Purification Kit (Promega, cat. # A1120) and susceptible isolates by PAP were sequenced using 400 Mbp long reads Illumina whole genome sequencing and heteroresistant isolates by PAP were sequenced with a combination of 600 Mbp long reads of nanopore sequencing and 400 Mbp long reads of Illumina whole genome sequencing. Raw paired-end sequence reads were analyzed using Bactopia pipeline version 3.0.1 (10)[1]. Briefly, the paired-end sequencing reads were fed into Bactopia pipeline with assigned quality checks, and reads below the quality requirements were filtered out. The remaining reads were assembled using Shovill version 1.1.0 (https://github.com/tseemann/shovill.git). Assembled contigs were annotated using prokka version 1.14.6 (11). To construct phylogenetic tree, Bactopia output files and raw reads were subjected to mashtree workflow on Bactopia. Mashtree workflow uses mash version 2.3 to create mash distance and estimate mutation rates between sequences and generates a newick format phylogenetic tree based on mash distance (12, 13). The phylogenetic tree was visualized and annotated using Interactive Tree of Life (iTOL) version 6 (14).

### Statistical Analyses

Statistical analyses were performed to determine associations between heteroresistance and overall, clinical, and microbiological outcomes. Descriptive and inferential statistics were performed for all variables. All categorical data were reported as percentages. Univariate analyses were conducted using Fisher’s Exact Test or Chi-square test for dichotomous variables and using Student’s t-test or Wilcoxon’s Rank Sum test for continuous variables. Multivariable analyses were conducted using logistic regression to evaluate the relationship between heteroresistance and treatment outcomes while addressing potential confounding. All p values were two-sided and a p value of < 0.05 was considered significant. A propensity score for the likelihood of a pathogen to be heteroresistant was developed by assessing univariate predictors of heteroresistance (p<0.20). These predictors were included in a logistic regression model predicting heteroresistance. The propensity score was calculated by adding together the β-coefficients of the model covariates. This propensity score was then included in logistic regression models assessing the association between heteroresistance and outcomes. Data analysis was performed using SAS software, version 9.4 (SAS Institute, Cary, NC).

## Results

### Bacterial Isolates and Patient Characteristics

Two-hundred and eighty-eight subjects with 291 isolates were included in the study. The 291 bacterial isolates that had an MIC below the CLSI resistance breakpoint for piperacillin/tazobactam as determined by BMD were tested for piperacillin/tazobactam heteroresistance by population analysis profile (PAP). Three subjects each had 2 UTI pathogens – in two of these instances both isolates were non-heteroresistant (and the subjects were categorized as non-heteroresistant) and in the other instance, one of the pathogens was heteroresistant and therefore the infection was categorized as heteroresistant. 33/288 (11.5%) of the patients were infected with piperacillin/tazobactam heteroresistant isolate and the remaining 255/288 (88.5%) were non-heteroresistant (Table 1). Overall, the patients infected by heteroresistant isolates had similar baseline characteristics as the patients infected by non-heteroresistant isolates except for patients with diabetes (Table 1). Interestingly, patients with diabetes had an increased likelihood of infection with a heteroresistant isolate (P-value 0.046) (Table 1).

**Table 1.** Characteristics of patients with non-heteroresistant and heteroresistant isolates. *Isolates that are susceptible by population analysis profile. ^a^ Ethnicity information is not available for a single patient in the heteroresistant category. ^b^ Baseline EGFRGR information is not available for 15 patients in the non-heteroresistant category and 2 in the heteroresistant. ^c^ Baseline CCINDEX information is not available for 2 patients in the non-heteroresistant category. ^d^ Enterobacterales baseline pathogens were genotyped for extended-spectrum beta-lactamases if their minimum inhibitory concentration was equal to or greater than 1 ug/mL or more for ceftazidime, ceftriaxone, cefepime, meropenem, or cefepime/enmetazobactam (8).

| Characteristic | Patients with a Non-heteroresistant Pathogen* (n=255) | Patients with a Heteroresistant Pathogen (n=33) | P Value |
| --- | --- | --- | --- |
| Age, Y |  |  |  |
| <65 | 170/255 (66.7%) | 21/33 (63.6%) |  |
| 65 to <75 | 52/255 (20.4%) | 9/33 (27.3%) |  |
| >=75 | 33/255 (12.9%) | 3/33 (9.1%) |  |
| Sex |  |  | 0.25 |
| Female | 161/255 (63.1%) | 17/33 (51.5%) |  |
| Race |  |  | 0.85 |
| Asian | 1/255 (.4%) | 0/33 (0%) |  |
| White | 242/255 (94.9%) | 32/33 (97%) |  |
| Other | 12/255 (4.7%) | 1/33 (3%) |  |
| Ethnicity<sup>a</sup> |  |  | 0.29 |
| Hispanic or Latino | 18/255 (7.1%) | 4/32 (12.5%) |  |
| Not Hispanic or Latino | 237/255 (92.9%) | 28/32 (87.5%) |  |
| BMI, mean (SD) | 26.0 (5.3) | 27.9 (6.6) | 0.12 |
| Baseline eGFR group, median (IQR), mL/min/1.732<sup>b</sup> | 62 (26) | 75 (34) | 0.34 |
| Infection Type |  |  | 0.16 |
| Acute pyelonephritis | 145/255 (58.9%) | 13/33 (39.4%) |  |
| Complicated UTI without removable source of infection | 64/255 (25.1%) | 12/33 (36.4%) |  |
| Complicated UTI with removable source of infection | 46/255 (18%) | 8/33 (24.2%) |  |
| Prior Antibiotic exposure |  |  | 0.10 |
| Short-acting antibiotic up to 24 h | 19/255 (7.5%) | 0/33 (0%) |  |
| Region |  |  | 0.76 |
| Eastern Europe | 193/255 (75.7%) | 25/33 (75.8%) |  |
| Americas | 15/255 (5.9%) | 1/33 (3%) |  |
| Other countries | 47/255 (18.4%) | 7/33 (21.2%) |  |
| Baseline Charlson Comorbidity Index Score<sup>c</sup> |  |  |  |
| CCINDEX <3 | 158/253 (62.5%) | 19/33 (57.6%) | 0.57 |
| Presence of concurrent bacteremia at baseline | 21/255 (8.2%) | 1/33 (3%) | 0.49 |
| Diabetes at baseline | 28/255 (11%) | 8/33 (24.2%) | **0.046** |
| Enterobacterales baseline pathogen | 246/255 (96.5%) | 29/33 (87.9%) | **0.049** |
| Enterobacterales baseline pathogen, extended-spectrum beta-lactamase encoding<sup>d</sup> | 37/246 (15%) | 18/29 (62.1%) | **<.0001** |

Extended spectrum beta-lactamase (ESBL)-encoding Enterobacterales were more likely to be heteroresistant when compared to non-ESBL-encoding Enterobacterales (P-value < 0.001) (Table 1). Although only 13/288 (4.5%) of patients were infected with non-Enterobacterales isolates, we observed that those isolates were more likely to be heteroresistant (4/13; 30.8%) than the Enterobacterales isolates (29/275; 10.5%) (P-value 0.049) (Table 1). The most common pathogen was *E. coli*, consisting of 216/291 (74.2%) of all isolates and 25/33 (75.8%) of the heteroresistant isolates, with the second most prevalent being *Klebsiella* spp. (30/291; 10.3%), consisting of 2/33 (6.1%) of the heteroresistant isolates (Supplementary Table 1).

### Heteroresistance and Treatment Outcome

The primary outcome of treatment failure at TOC occurred in 17/33 (51.5%) of the patients infected with heteroresistant isolates and 85/255 (33.3%) of those infected with non-heteroresistant isolates (Table 2), demonstrating that patients infected with a piperacillin/tazobactam heteroresistant isolate had a significantly increased failure rate (2.13 OR, 95% CI 1.02, 4.41) (Table 2). In adjusted analysis, after controlling for the propensity for heteroresistance (propensity score includes acute pyelonephritis infection, BMI, and diabetes), heteroresistance was associated with treatment failure with a similar effect size, although this did not reach statistical significance (OR 1.74, 95% CI 0.82, 3.71) (Table 2). However, subgroup analysis revealed that patients with a cUTI without a removable source had a significantly higher rate of treatment failure (OR 4.68, 95% CI 1.15, 18.97), and that this remained significant in adjusted analysis controlling for the propensity for heteroresistance (OR 4.56, 1.12, 18.62) (Table 3, Supplementary Table 3).

**Table 2.** Treatment outcomes of patients infected with non-heteroresistant or heteroresistant isolates. *Isolates that are susceptible by population analysis profile. † Adjusted for propensity score based on acute pyelonephritis infection, BMI, and diabetes

| Outcome | Non-heteroresistant<br>* n=255 | Heteroresistant<br>n=33 | Crude Odds Ratio<br>(95% CI) | Adjusted Odds Ratio †<br>(95% CI) |
| --- | --- | --- | --- | --- |
| Overall Failure | 85 (33.3%) | 17 (51.5%) | 2.13 (1.02, 4.41) | 1.74 (0.82, 3.70) |
| Microbiological Failure | 79 (31%) | 16 (48.5%) | 2.10 (1.01, 4.36) | 1.70 (0.80, 3.64) |
| Clinical Failure | 10 (3.9%) | 3 (9.1%) | 2.45 (0.64, 9.40) | 1.18 (0.35, 3.99) |

**Table 3.**
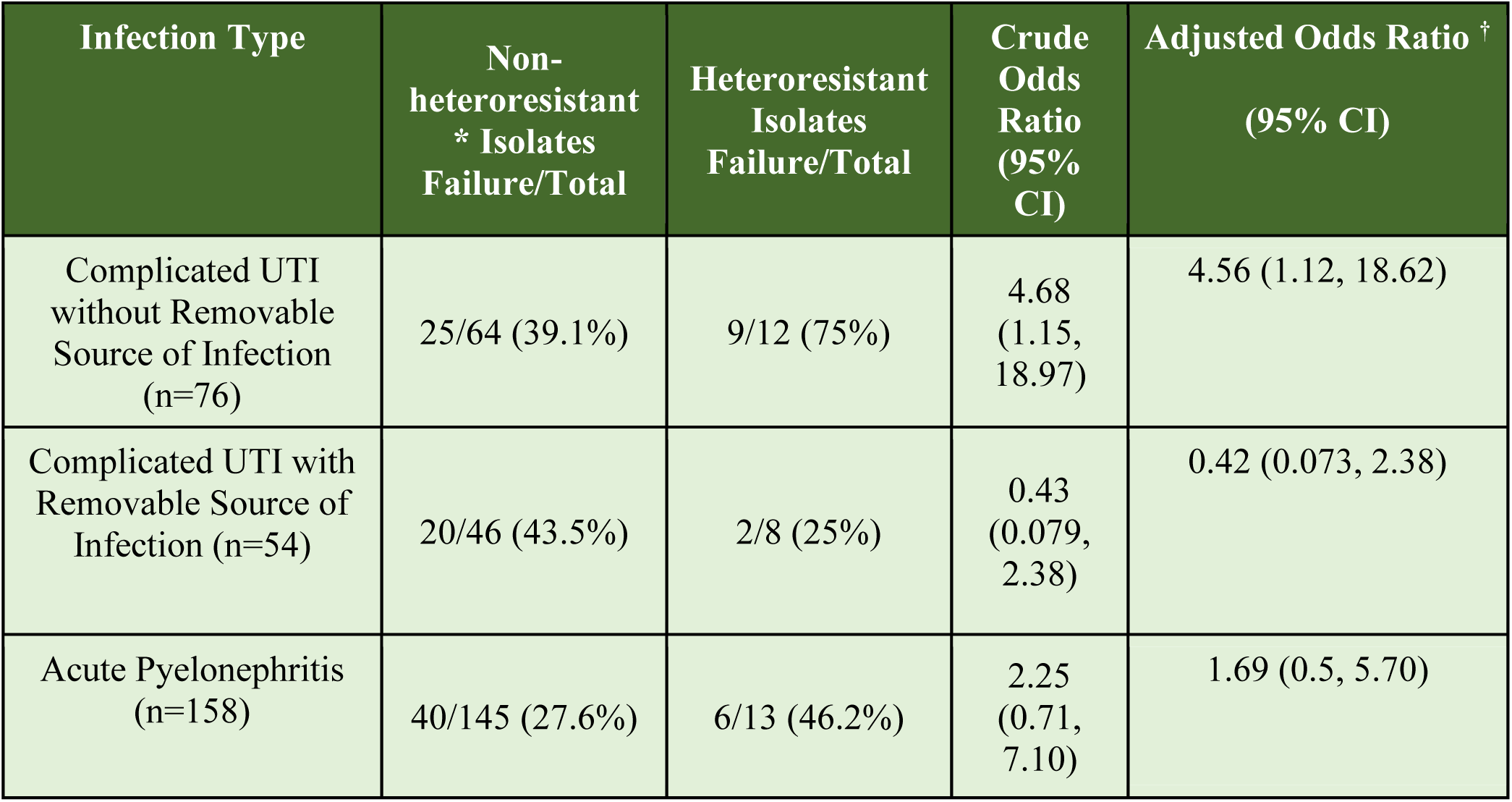
Association of heteroresistance and treatment failure stratified by infection type. *Isolates that are susceptible by population analysis profiles. †Adjusted for propensity score based on acute pyelonephritis infection, BMI, and diabetes

In addition to heteroresistance being associated with an increase in overall treatment failure, we observed that heteroresistance was associated with a significant increase in microbiological failure (OR 2.10, 95% CI 1.01, 4.36) (Table 2). After controlling for the propensity for HR, while the association of heteroresistance with microbiologic failure had a similar effect size, the association no longer reached statistical significance (OR 1.70, 95% CI 0.80, 3.64) (Table 2). Patients infected with heteroresistant isolates had a higher frequency of clinical failure compared to those infected with non-heteroresistant isolates, although this did not reach statistical significance in the unadjusted (OR 2.45, 95% CI 0.64, 9.40) or adjusted analysis controlling for the propensity for HR (OR 1.18, 95% CI 0.35, 3.99) (Table 2).

Interestingly, heteroresistant isolates were more likely to have an elevated MIC compared to the non-heteroresistant isolates, with 36.4% of heteroresistant isolates having an MIC of 16/4 ug/mL or 32/4 ug/mL compared to 1.9% of non-heteroresistant isolates (OR 28.9, 95% CI 9.3, 89.9, Supplementary Figure 2). Furthermore, there was an observable trend that treatment failure increased as MIC increased amongst susceptible isolates. Interestingly, this trend was not observed amongst the heteroresistant isolates (Supplementary Table 2).

## Discussion

Our study aimed to assess the potential contribution of heteroresistance to antibiotic treatment failure. The clinical trial data and isolates used allowed us to examine this question due to the large size of the patient cohort, regimented monotherapy treatment protocol, and uniform assessment of treatment outcomes (8). We observed that patients infected with piperacillin/tazobactam heteroresistant isolates had a significant, greater than two-fold higher rate of treatment failure when compared to patients infected with non-heteroresistant isolates (Table 2) and greater than four-fold higher rate of treatment failure among patients without a removable source of infection (Table 3).

Previous research revealed that the resistant subpopulations in heteroresistant isolates can rapidly replicate during antibiotic exposure, leading to their enrichment (3, 15). These findings suggested that heteroresistant isolates might drive treatment failure. Following this hypothesis, results from murine infection models demonstrated that heteroresistance can indeed cause treatment failure (3–5). This convergence of evidence, combined with the results presented here, demonstrates that the presence of heteroresistance increases the likelihood of treatment failure.

The MIC for every isolate included in this study was below the resistance breakpoint for piperacillin/tazobactam when tested by broth microdilution (BMD) in accordance with 2024 CLSI standards. The increase in treatment failure rates exhibited by heteroresistant isolates, juxtaposed with their apparent susceptibility according to BMD, emphasizes the capacity of heteroresistant isolates to remain undetected by conventional susceptibility testing while concurrently causing a higher rate of treatment failure. The most likely reason that BMD is unable to detect the piperacillin/tazobactam resistant subpopulation in heteroresistant isolates is that these cells are too infrequent in the overall population (16). BMD utilizes an inoculum of ∼5×10^4^ CFU per well. If a heteroresistant isolate harbors 1 in 100,000 resistant cells, a resistant cell may not be present in the BMD inoculum. On average, the heteroresistant isolates in this study had less than 1 in 100,000 resistant cells surviving at the breakpoint concentration of piperacillin/tazobactam (Supplementary Figure 3).

These population dynamics may also explain the higher rate of piperacillin/tazobactam treatment failure among patients without a removable catheter and infected with heteroresistant isolates. Given that the average frequency of resistant cells was 1 in 100,000 among heteroresistant isolates in this study, it follows that a high burden of total infecting bacterial cells would be necessary for sufficient resistant cells to be present to withstand piperacillin/tazobactam treatment and remain to cause treatment failure. Having a non-removable source provides a surface on which total bacterial load can persist, and in the presence of active therapy, a resistant subpopulation is present in sufficient number to ultimately mediate treatment failure.

We noted and adjusted for (via a propensity score) three patient characteristics that were associated with the patient being infected by a heteroresistant isolate: acute pyelonephritis, BMI, and diabetes status. Potential reasons why these variables were associated with heteroresistance are described in Supplementary Figure 4. These three variables were also confounders of the association between heteroresistance and treatment failure (Supplementary Figure 4). Furthermore, we observed a strong association between ESBL-encoding Enterobacterales isolates and heteroresistance (Table 1). Previous studies have demonstrated that heteroresistant strains often harbor resistant subpopulations generated via increased copy number of specific genomic regions encoding antibiotic resistance genes such as beta-lactamases (17–20). The higher prevalence of ESBL-encoding isolates within the heteroresistant cohort could therefore signify their tendency to amplify ESBL genes and thus generate resistant subpopulations. This puts ESBL production on the causal pathway between heteroresistance and treatment failure and accordingly was not adjusted for in the propensity score (Supplementary Figure 4). To further analyze this correlation, non-heteroresistant and heteroresistant *E. coli* isolates were sequenced and a phylogenetic tree was generated (Supplementary Figure 5). We found that ESBL-encoding *E. coli* were often clustered together. Additionally, we found a strong clustering of *E. coli* heteroresistant isolates within the ESBL-encoding cluster (Supplementary Figure 5). Further studies will need to be conducted to better understand these observed phylogenetic clusters. We also observed an association between increased prevalence of heteroresistance in non-Enterobacterales compared to Enterobacterales, although the explanation for this finding is unclear. To further meaningfully explore this association, a much larger sample size of non-Enterobacterales isolates would be required.

When treating a patient for a serious infection, there are multiple concurrent factors that influence the ultimate treatment outcome – treatment factors (e.g., antibiotic potency, dosing, penetration to the site of infection, etc.), host factors (e.g., immune status, baseline comorbidities, etc.), and pathogen/infection factors (e.g., virulence, extent of organism load, etc). Antibiotic susceptibility testing provides crucial information to determine if an organism is susceptible to various antibiotics. While ‘susceptible’ generally equates to clinical efficacy of an antibiotic against the isolated bacterium, treatment failure can nonetheless occur. Understanding factors that mediate treatment failure despite apparent antibiotic susceptibility as measured by current clinical diagnostic tests is important to improve patient outcomes. The data presented here demonstrate that heteroresistance is a contributor to treatment failure, at least in the setting evaluated here. Further quantifying the prevalence of this phenotype across bacterial species and infection types will be necessary in future studies. Not only do the current findings highlight the need for better understanding of antibiotic treatment strategies to overcome heteroresistance, but advancement of novel susceptibility testing methods capable of detecting heteroresistance should be a focus as well.

This study has several limitations to note. First, the sample size for patients infected with heteroresistant isolates was relatively small, which in multivariable analysis, limited the ability to demonstrate a significant association between heteroresistance and treatment failure, and also limited the power to detect associations between heteroresistance and outcomes in sub-group analyses. Future studies with larger cohorts of patients infected with heteroresistant isolates can address this limitation. Second, as these isolates originated from a previous clinical trial with strict treatment guidelines, our findings do not necessarily reflect the impact of heteroresistance during “real world” use of antibiotics. Third, these isolates were from patients with cUTI and AP and therefore may not be reflective of other bacterial infection types. Despite these limitations, the strengths of this study include the uniform antibiotic treatment and patient outcome assessment as well as the strong association between heteroresistance and treatment failure.

In conclusion, we show that patients infected with piperacillin/tazobactam heteroresistant isolates have an increased likelihood of treatment failure when compared to those infected with non-heteroresistant isolates. Heteroresistance, particularly among Gram-negative pathogens, is an important and often overlooked phenomenon that may play a crucial role in antibiotic treatment failure.

## Data Availability

All data produced in the present study are available upon reasonable request to the authors

## Supplementary Materials

**Supplementary Table 1.** Treatment failure rate as a function of heteroresistance for each pathogen in the study. *Isolates that are susceptible according to population analysis profiles.

| Species | Non-heteroresistant* isolates/Total Isolates | Heteroresistant isolates/Total isolates | Non-heteroresistant* Failure Rate | Heteroresistant Failure Rate |
| --- | --- | --- | --- | --- |
| TOTAL | 258/291 (88.7%) | 33/291 (11.3%) | 86/258 (33.3%) | 17/33 (51.5%) |
| Enterobacterales | 249/278 (89.6%) | 29/278 (10.4%) | 80/249 (32.1%) | 15/29 (51.7%) |
| *Escherichia* | 191/216 (88.4%) | 25/216 (11.6%) | 59/191 (30.9%) | 12/25 (48%) |
| *Enterobacter* | 3/4 (75%) | 1/4 (25%) | 1/3 (33.3%) | 1/1 (100%) |
| *Citrobacter* | 3/3 (100%) | 0/3 (0%) | 0/3 (0%) | N/A |
| *Klebsiella* | 28/30 (93.3%) | 2/30 (6.7%) | 8/28 (28.6%) | 2/2 (100%) |
| *Morganella* | 1/1 (100%) | 0/1 (0%) | 0/1 (0%) | N/A |
| *Proteus* | 19/20 (95%) | 1/20 (5%) | 8/19 (42.1%) | 0/1 (0%) |
| *Providencia* | 1/1 (100%) | 0/1 (0%) | 1/1 (100%) | N/A |
| *Serratia* | 3/3 (100%) | 0/3 (0%) | 3/3 (100%) | N/A |
| Non-Enterobacterales | 9/13 (69.2%) | 4/13 (30.8%) | 6/9 (66.7%) | 2/4 (50%) |
| *Acinetobacter* | 2/2 (100%) | 0/2 (0%) | 1/2 (50%) | N/A |
| *Pseudomonas* | 7/11 (63.6%) | 4/11 (36.4%) | 5/7 (71.4%) | 2/4 (50%) |

**Supplementary Table 2.**
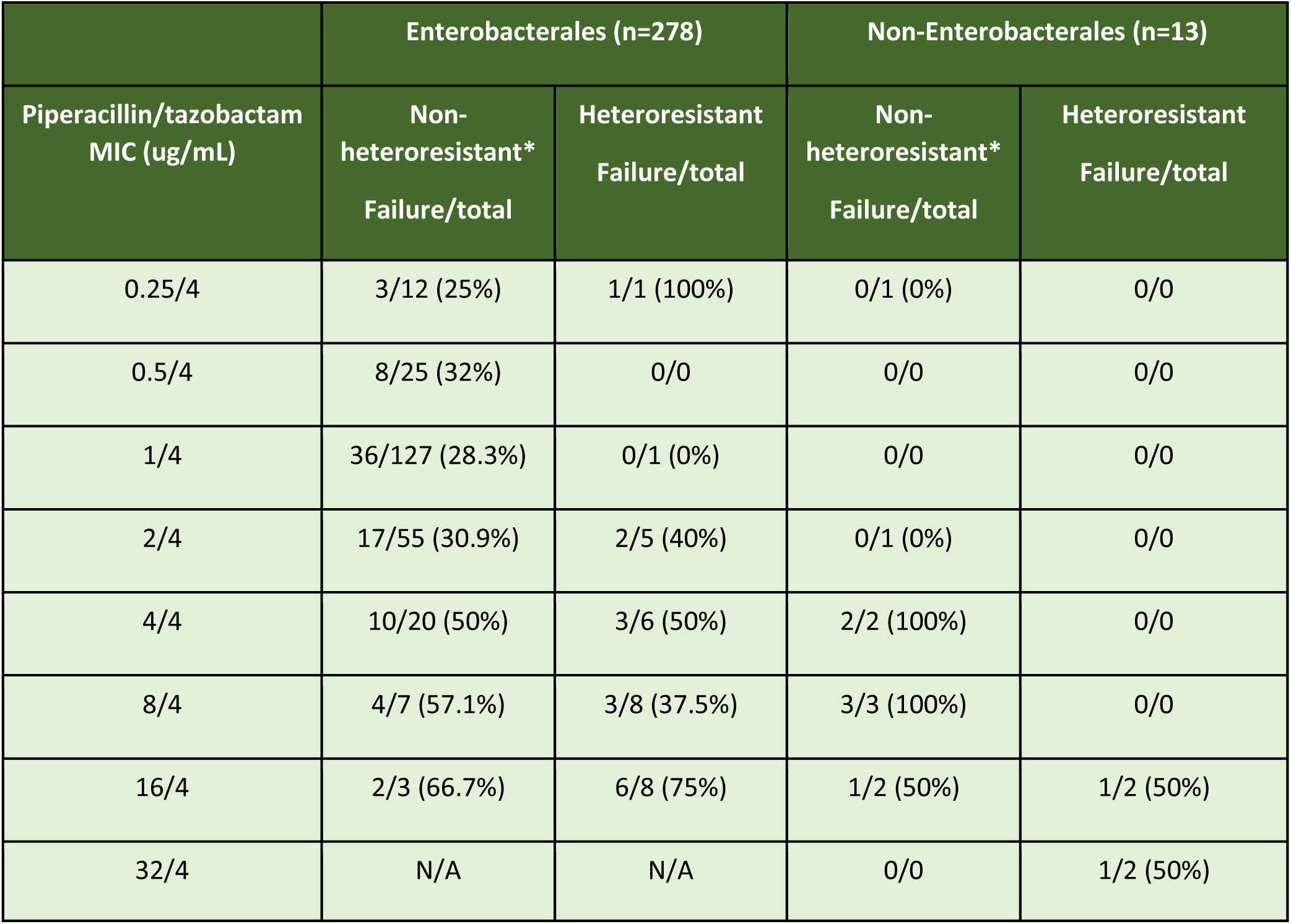
Treatment failure rates by piperacillin/tazobactam MIC for Enterobacterales and non-Enterobacterales. *Isolates that are susceptible by population analysis profiles.

|  | Enterobacterales (n=278) |  | Non-Enterobacterales (n=13) |  |
| --- | --- | --- | --- | --- |
| Piperacillin/tazobactam<br>MIC (ug/mL) | Non-heteroresistant*<br>Failure/total | Heteroresistant<br>Failure/total | Non-heteroresistant*<br>Failure/total | Heteroresistant<br>Failure/total |
| 0.25/4 | 3/12 (25%) | 1/1 (100%) | 0/1 (0%) | 0/0 |
| 0.5/4 | 8/25 (32%) | 0/0 | 0/0 | 0/0 |
| 1/4 | 36/127 (28.3%) | 0/1 (0%) | 0/0 | 0/0 |
| 2/4 | 17/55 (30.9%) | 2/5 (40%) | 0/1 (0%) | 0/0 |
| 4/4 | 10/20 (50%) | 3/6 (50%) | 2/2 (100%) | 0/0 |
| 8/4 | 4/7 (57.1%) | 3/8 (37.5%) | 3/3 (100%) | 0/0 |
| 16/4 | 2/3 (66.7%) | 6/8 (75%) | 1/2 (50%) | 1/2 (50%) |
| 32/4 | N/A | N/A | 0/0 | 1/2 (50%) |

**Supplementary Table 3.** Characteristics of patients without a removable source and either non-heteroresistant or heteroresistant isolates. *Isolates that are susceptible by population analysis profile. ^a^ Ethnicity information is not available for a single patient in the heteroresistant category. ^b^ Baseline EGFRGR information is not available for 15 patients in the non-heteroresistant category and 2 in the heteroresistant. ^c^ Baseline CCINDEX information is not available for 2 patients in the non-heteroresistant category. ^d^ Enterobacterales baseline pathogens were genotyped for extended-spectrum beta-lactamases if their minimum inhibitory concentration was equal to or greater than 1 ug/mL or more for ceftazidime, ceftriaxone, cefepime, meropenem, or cefepime/enmetazobactam.

| Characteristic | Non-heteroresistant*<br>(n=64) | Heteroresistant<br>(n=12) | P Value |
| --- | --- | --- | --- |
| Age, Y |  |  | 0.62 |
| <65 | 19/64 (29.7%) | 4/12 (33.3%) |  |
| 65 to <75 | 32/64 (50%) | 7/12 (58.3%) |  |
| >=75 | 13/64 (20.3%) | 1/12 (8.3%) |  |
| Sex |  |  | 0.53 |
| Female | 23/64 (35.9%) | 3/12 (25%) |  |
| Race |  |  | 0.63 |
| White | 62/64 (96.9%) | 12/12 (100%) |  |
| Other | 2/64 (3.1%) | 0/12 (0%) |  |
| Ethnicity<sup>a</sup> |  |  | 1.00 |
| Hispanic or Latino | 2/64 (3.1%) | 0/11 (0%) |  |
| Not Hispanic or Latino | 62/64 (96.9%) | 11/11 (91.7%) |  |
| BMI, mean (SD) | 28.6 (4.9) | 29.8 (6.9) | 0.60 |
| Baseline eGFR group, median (IQR),<br>mL/min/1.732<sup>b</sup> | 63 (23) | 72.5 (32.5) | 0.33 |
| Prior Antibiotic exposure |  |  | 0.53 |
| Short-acting antibiotic up to 24h | 2/64 (3.1%) | 0/12 (0%) |  |
| Region |  |  | 0.82 |
| Eastern Europe | 46/64 (71.9%) | 9/12 (75%) |  |
| Americas | 2/64 (3.1%) | 0/12 (%) |  |
| Other countries | 16/64 (25%) | 3/12 (25%) |  |
| Baseline Charlson Comorbidity Index Score<sup>c</sup> |  |  | 0.76 |
| CCINDEX <3 | 28/64 (44.4%) | 6/12 (50%) |  |
| Presence of concurrent bacteremia at baseline | 4/64 (6.3%) | 0/12 (0%) | 1.00 |
| Diabetes at baseline | 12/64 (18.8%) | 3/12 (25%) | 0.69 |
| Enterobacterales baseline pathogen | 62/64 (96.9%) | 10/12 (83.3%) | 0.12 |
| Enterobacterales baseline pathogen, extended-spectrum beta-lactamase encoding<sup>d</sup> | 9/62 (14.5%) | 4/12 (40%) | 0.07 |

**Supplementary Figure 1.**
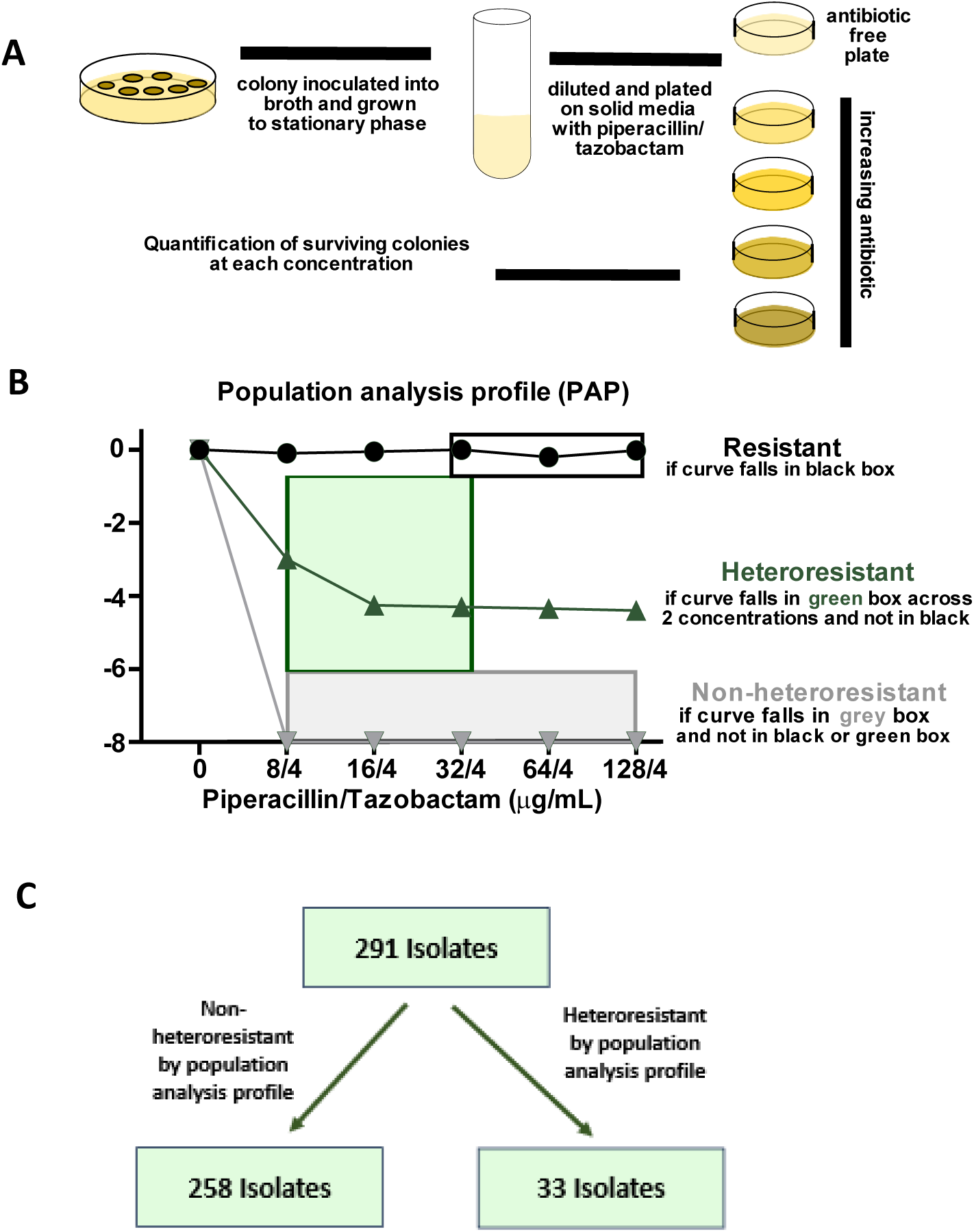
Population Analysis Profile. (A) A clinical isolate is streaked for isolation, inoculated into broth, and incubated overnight. The following day the bacterial culture is serially diluted and plated on plates with either no antibiotic or increasing concentrations of piperacillin/tazobactam. (B) After overnight incubation, the colonies are enumerated and the number of surviving colonies on the various drug concentrations are compared to the number on the plate not containing drug. (C) Isolates were designated either non-heteroresistant (susceptible) or heteroresistant by population analysis profiles. Adopted from Choby et al, Lancet Infect Dis. 2021;21(5):597-8.

**Supplementary Figure 2.**
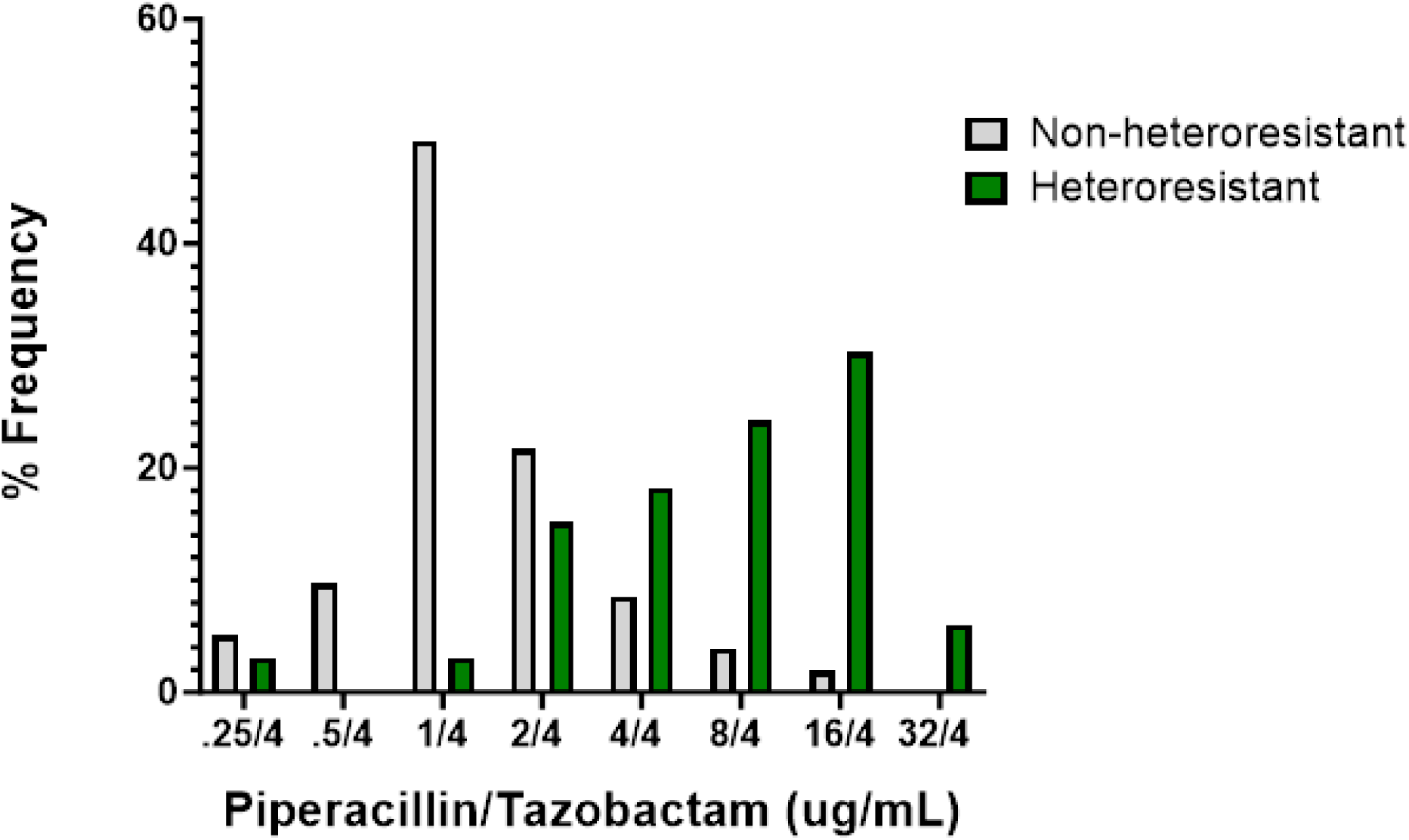
Frequency of isolates with each piperacillin/tazobactam MIC based on broth microdilution testing for non-heteroresistant and heteroresistant isolates.

**Supplementary Figure 3.**
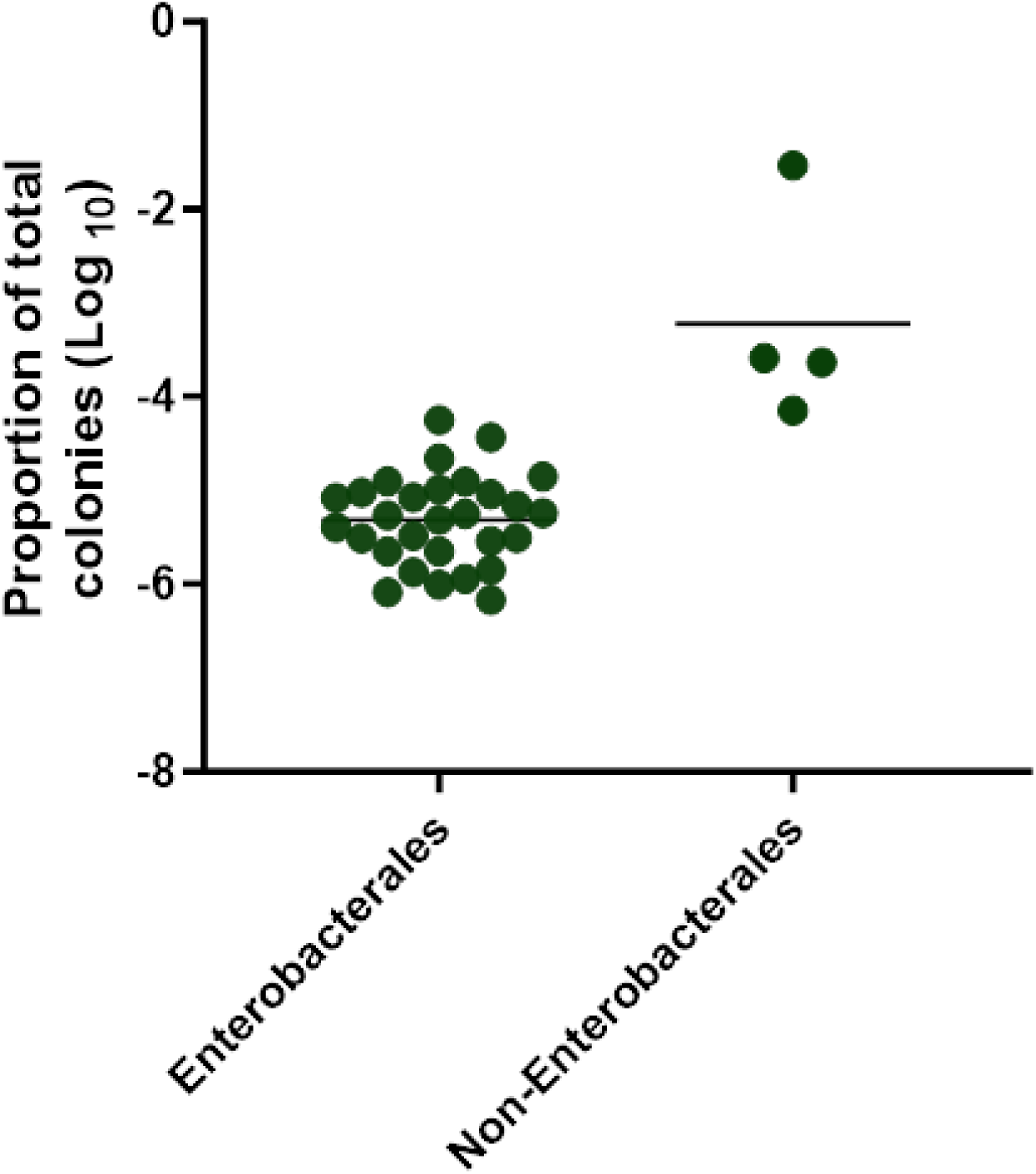
Frequency of the resistant subpopulation in heteroresistant isolates. Piperacillin/tazobactam heteroresistant Enterobacterales and non-Enterobacterales isolates were plated on 32/4ug/mL piperacillin/tazobactam and the proportion of surviving cells is shown relative to agar without antibiotic.

**Supplementary Figure 4.**
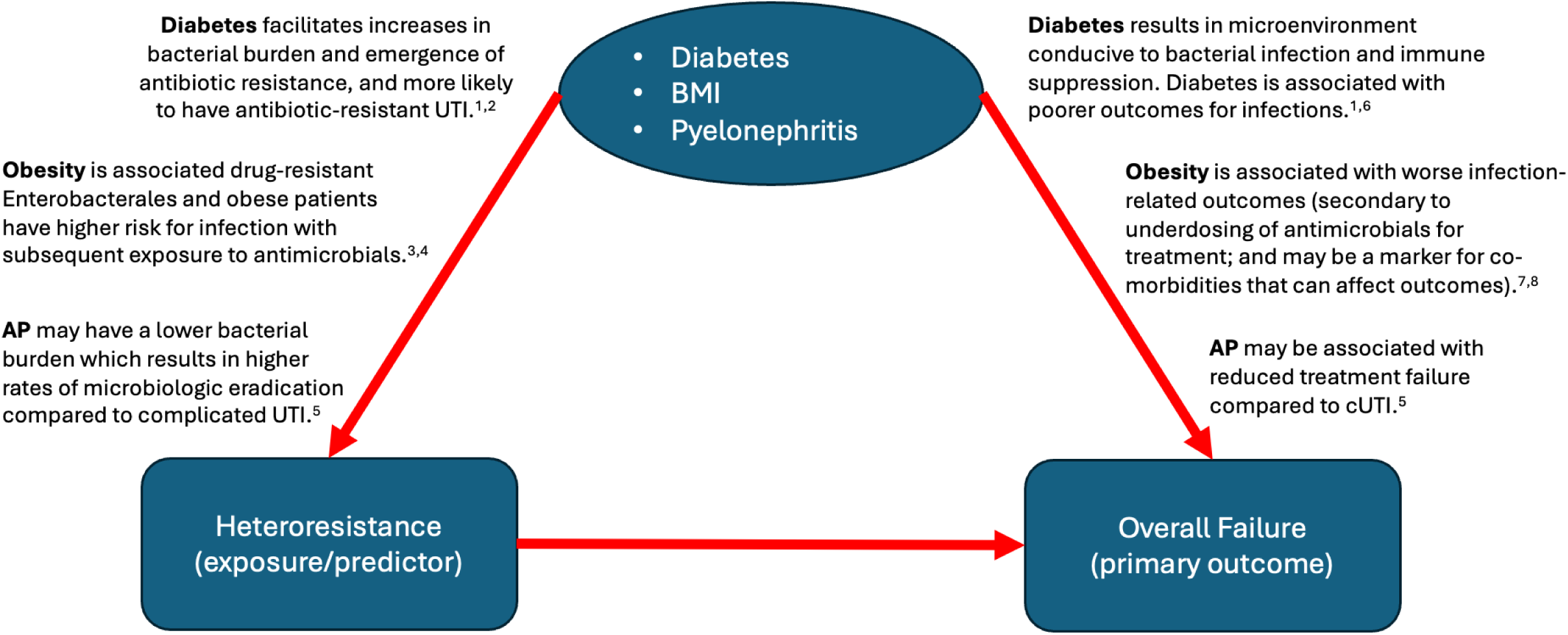
Directed acyclic graph (DAG) indicating potential reasons why variables were associated with heteroresistance.

**Supplementary Figure 5.**
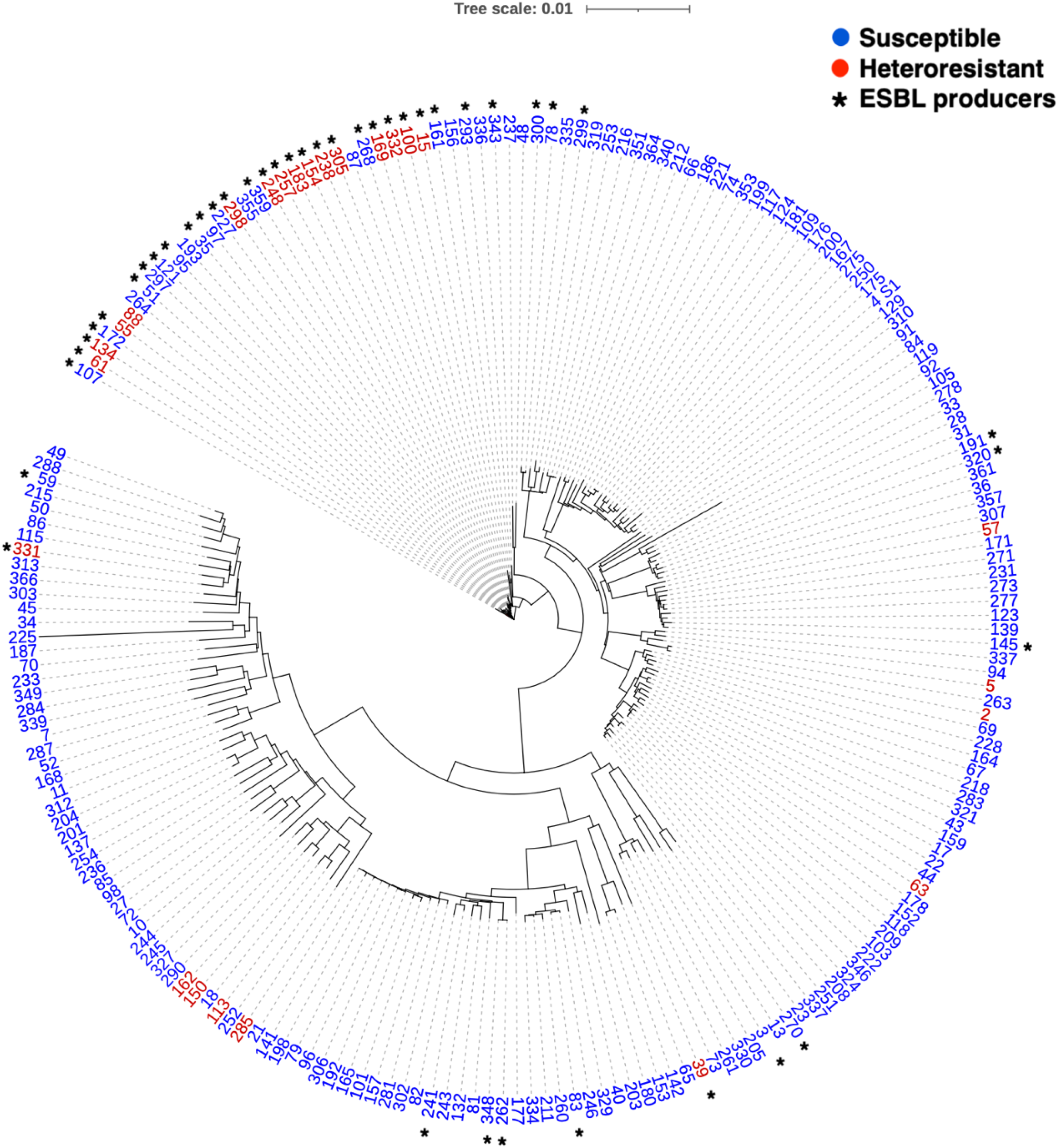
Phylogenetic clustering of heteroresistant, ESBL producers among *E. coli* in the study. Genome sequences for *E. coli* isolates in the study were phylogenetically clustered. Genomes for non-heteroresistant isolates (susceptible by population analysis profile testing; blue) and heteroresistant isolates (red) are indicated. ESBL isolates are indicated with an asterisk.

